# Examining the role of systemic inflammation as a mediator of the glycaemia-brain volume associations in women

**DOI:** 10.1101/2025.08.01.25332698

**Authors:** Nasri Fatih, Nish Chaturvedi, Victoria Garfield, Carole H. Sudre, Richard J Silverwood, David M Cash, Ian B Malone, Josephine Barnes, Marcus Richards, Jonathan M Schott, Alun D Hughes, Sarah-Naomi James

**Affiliations:** Nuffield Department of Population Health, Big Data Institute, University of Oxford, Oxford, UK; Unit for Lifelong Health and Ageing at UCL, London, UK; Dementia Research Centre, UCL Queen Square Institute of Neurology, University College London, London, UK; Centre for Longitudinal Studies, UCL Social Research Institute, University College London, London, UK; UK Dementia Research Institute at UCL, University College London, London, UK; Department of Pharmacology and Therapeutics, University of Liverpool. Ashton Street University of Liverpool, L69 3GE

## Abstract

Previous studies have found that diabetes and its mechanistic factors (e.g. glycaemia) are associated with poorer cognitive and brain health. There is also growing evidence of sex differences in how diabetes manifests itself and impacts the brain. The mechanisms through which this association manifests itself are still poorly understood, but the possible role of inflammation has been proposed. This study aims to explore whether the relationship between mid-life glycaemia and brain volumes in later-life in women is mediated by systemic inflammation. The sample consisted of female participants from the National Survey of Health and Development (NSHD) who underwent neuroimaging as part of the Insight 46 sub-study. Path analysis models were then constructed between glycaemic markers (age 60-64) and brain health outcomes (age 69-71) with adjustments for social and metabolic confounders (age 60-64). Although glycaemia was mostly associated with a higher systemic inflammatory state in two of the three markers (e.g., HbA1c and interleukin-6: β = 0.05 [0.02. 0.01], p = 0.001 and glycoprotein A: β = 0.02 [=-0.01. 0.02], p = 0.001), we did not find a relationship between inflammation and our brain volume markers [whole brain, grey matter and white matter] (e.g. interleukin-6 and whole brain volume: β = -3.1 [=-7.7. 1.5], p = 0.2; interleukin-6 and grey matter: β = -0.3 [=-1.8. 1.2], p = 0.7), thus no mediated effect between the glycaemic markers and outcomes via the pathway of systematic inflammation. This raises the possibility alternative mechanistic pathway, to inflammation, playing a role in the relationship between hyperglycaemia and brain health outcomes.

## Introduction

Previous studies have shown an association between diabetes and poorer cognitive health as well as higher risk of dementia. In our previous work, we found that mid-life glycaemia (as indexed by HbA1c) was associated with poorer brain health later in life exclusively in women. (Fatih et al., 2022). This aligns with other studies have also reported An important next step that follows from these findings is to investigate potential mechanistic explanations for these sex differences. One potential mediating factor of this relationship could be inflammation since there is growing evidence of: 1) evidence that inflammation is a central feature of type 2 diabetes (T2D)(Gupta et al., 2023) and 2) associations between systemic inflammatory markers such as interleukin-6 (IL-6) and brain health outcomes both in animal models and humans (Marsland et al., 2008; Singh-Manoux et al., 2012; Walker et al., 2017) 3) higher systemic inflammation in women (Cartier et al., 2009). Sex differences in inflammation levels may particularly become more prominent and influential following the decline of oestrogen’s neuroprotective and anti-inflammatory effect during the perimenopause and following the menopause (Cioffi et al., 2002; Su & Freeman, 2009). A sex-specific transcriptomic analysis also revealed that female microglia exhibit evidence of both higher levels of inflammation and Alzheimer’s disease-related (AD) genes compared to male microglia suggesting a higher neuroinflammatory response in women, which may have broader implications for sex differences in brain health and disease (Coales et al., 2022). Thus, systemic inflammation may mediate the sex-specific associations between glycaemia, and volumetric brain health predominantly observed in women.

Here we expand on our previous findings and investigate the extent through which the relationship between glycaemic markers (HbA_1c_ and glucose) at age 60-64 and volumetric brain measures at age ∼70 in female participants was mediated by systemic inflammation. Inflammation was indexed by IL-6, C-reactive protein (CRP) and GlycA, a spectroscopic marker of systemic inflammation related to the levels and degree of glycosylation of various acute phase proteins.

## Materials and Methods

### Ethical approval, consent to participate and data access

The study protocol—including this secondary analysis of NSHD data—was reviewed and approved *before any data were accessed* by the London-Queen Square NHS Research Ethics Committee (14/LO/1073) and by the Scotland A Research Ethics Committee (14/SS/1009). All NSHD waves obtain written informed consent from participants in accordance with the Declaration of Helsinki. The dataset supplied to the authors for the present analysis was fully pseudonymised; direct identifiers are retained on a separate, access-controlled server. The lead author first accessed the anonymised extract for research purposes on 3 March 2022.

### Sample

The sample consisted of participants of the National Survey of Health and Development (NSHD). NSHD is a British birth cohort originally made up of 5,362 boys and girls born across mainland Britain during the same week in 1946 (Kuh et al., 2016). In 2006, the study members (aged 60-64 at the time) received postal questionnaires and were invited to attend a clinic visit, when blood samples were taken to index glycaemic health and systemic inflammation. Between 2015-2018, a subset of 502 NSHD participants were enrolled into the Insight 46 sub-study to undergo neuroimaging and further assessments. Selection was restricted to those who had previously attended the clinic-based assessment at age 60–64, had previously intimated they were willing to attend a clinic visit in London and for whom relevant data in childhood and adulthood are available (Lane et al., 2017a).

### Investigations

#### Exposure variables

HbA_1c_ and glucose were measured in a fasting blood sample collected at age 60–64. HbA_1c_ was measured by ion exchange High-performance liquid chromatography (HPLC) on a Tosoh analyzer (Tosoh Bioscience, Tessenderlo, Belgium) whereas glucose was measured via an enzymatic assay using hexokinase coupled to glucose 6-phosphate dehydrogenase on a Siemens Dimension Xpand analyzer (Siemens Medical Solutions, Erlangen, Germany).

#### Systemic inflammation mediators

Inflammatory markers were measured from fasting blood samples collected at age 60–64 during clinic or home visits. Interleukin 6 (IL-6)was assayed from serum using ELISA (inter-assay CV: 6.5%) and reported in pg/L. CRP was measured using a high-sensitivity particle-enhanced immunoturbidimetric assay (CV: 6.28%, detection limit: 1 ng/ml) and reported in mg/L. Glycoprotein acetyls (GlycA), specifically alpha1-acid glycoprotein, were analyzed via high-throughput nuclear magnetic resonance (NMR) metabolomics on serum samples (no freeze-thaw cycles) using Bruker spectrometers, with most inter-assay CVs below 5%, as detailed in Soininen et al. (2015).

#### Confounders

Confounders were identified based on prior knowledge of associations between hyperglycaemia and inflammation, and inflammation and brain health, which were then represented through a directed acyclic graph (DAG) (see supplementary figure 1B).

Confounders were considered for both the exposure-mediator relationships and the mediator-outcome relationships. In this analysis, the confounders considered were: **Socioeconomic position:** Childhood socioeconomic position (SEP) was measured as father’s occupational social class recorded at age 4 (or if missing, at age 11) and categorised into manual or non-manual according to the UK Registrar General’s Standard’s Occupation Classification. Adult SEP was based on head of household occupation at age 53 years and categorised into manual or non-manual.

##### Education

The highest educational attainment or training qualification achieved by 26 years was classified according to the Burnham scale (Hatch et al., 2009) and grouped into the following: no qualification; below ordinary secondary qualifications (e.g., vocational qualifications); ordinary level qualifications (‘O’ levels or their training equivalents); advanced level qualifications (‘A’ levels or their equivalents); or higher education (degree or equivalent).

##### Alcohol

Information on alcohol consumption over the previous 7 days was obtained by a self-completed questionnaire completed between the ages of 60-64. Responses were totalled to provide an approximate measure of drinks per week, where a drink (or unit in UK terminology) contains ∼9.0 g of alcohol. Participants were then dichotomised into two categories: those who drunk under 14 units of alcohol per week and those who drunk 14 or more units of alcohol per week.

##### Smoking

Smoking status at age 60-64 was assessed by self-report and was classified into three groups: current smokers, ex-smokers, and never-smokers.

##### Physical activity

Physical activity information was collected at age 60–64 using the EPIC physical Activity questionnaire-2 (Richardson et al., 1994). This assessed how often participants had had taken part in any sports, vigorous leisure activities or exercise in the previous 4 weeks. Responses were categorised into: 1) not active (no participation in physical activity/month), 2) moderately active (participated 1–4 times/month) and 3) most active (participated 5 or more times/month).(Black et al., 2015)

##### Body mass index

body mass index was calculated using the following equation: (weight(kg)/height(m^2^)). Height and weight (at age 60-64) were measured by nurses using standardised protocols.

##### Arthritis

Arthritis status was assessed through questionnaires asking whether participants had taken non-steroidal anti-inflammatory medication at age 60-64 or they had been diagnosed with the condition.

##### COPD

Chronic obstructive pulmonary disease (COPD) at age 60-64 was identified based on the presence of airflow obstruction defined by the ratio of FEV_1_/FVC of less than the lower limit of normal or 0.7.(Torén et al., 2021).

#### Volumetric neuroimaging outcomes

We ascertained metrics of whole brain volume (WBV), grey matter (GM) and white matter (WM) volumes using validated pipelines. In brief, individuals underwent MRI scanning on the same Biograph mMR 3T PET-MRI scanner (Siemens Healthcare, Erlangen), which included high resolution 3D (1·1 mm isotropic) T1-weighted and T2-weighted Fluid attenuated inversion recovery (FLAIR) sequences as previously described (Lane et al., 2017; Murray-Smith et al., 2024).

Following closely the pipelines detailed in Eshagi and colleagues (Eshaghi et al., 2019), original parcellation applying GIF (Cardoso et al., 2015)was used to identify WMH (BaMoS). These lesion maps were used to inpaint the T1 weighted (Prados et al., 2016) image with healthy looking tissue before performing the parcellation on the healthy looking image to avoid any possible processing bias introduced by the lesions. WBV was segmented using Multi-Atlas Propagation and Segmentation (MAPS). (Leung et al., 2011)

### Statistical analyses

To address the extent to which systemic inflammation mediates the relationship between glycemia and brain volume, we conducted a two-component mediation analysis using the ‘sem’ package (StataCorp) in Stata 17 (StataCorp, College Station, TX, USA). This approach, which is analogous to path analysis, decomposes the total ‘effect’ (i.e., association between exposure and outcome) into a direct, and an indirect (or mediated) effect (VanderWeele, 2011, 2013) [see supplementary figure 2]. While a three-component mediation analysis which also included a mediated interactive effect was contemplated, for reasons that will become apparent from the results, this was not performed. Note that in this analysis framework the term “effect” is used to describe the relationships between exposure, mediator and outcome. Such relationships or associations can only be interpreted as causal under very strong assumptions that are arguably never (or perhaps almost never) satisfied. Some eschew the term ‘effect’ in all epidemiological studies for this reason, but we emphasise that casual interpretations should not be applied to the term “effect” in this context. This is discussed by Hernan and colleagues (Cole & Hernán, 2002; Hernán, 2018)

All models were estimated using full information maximum likelihood (FIML), which assumes linear relationships and multivariate normality but allows for missing data under a missing at random (MAR) assumption and was therefore considered preferable to estimation based on complete case data. As a sensitivity analysis, we also conducted a complete case analysis using an estimator assuming multivariate normality to assess whether violations of normality due to categorical or missing data handling in FIML affected our results.

Due to their skewed distribution, IL-6 and CRP were log-transformed to conform with the multivariate normal assumptions of structural equation modelling. In addition, participants with CRP values below the limit of detection (LOD), (i.e., under the value of 1mg/L) were assigned a value of 1 mg/L.

The path analysis was conducted to model the relationships between the glycaemic markers and each individual inflammatory marker (IL-6, CRP and GlycA) and outcome (WBV, WM volume and GM volume). This model was initially created as a minimally adjusted model (considered age, TIV) [see Supplementary figure 1A] but then built into a fully confounder-adjusted model including child and adult SEP, education, BMI, alcohol, smoking status, physical activity, arthritis medication use, and COPD, and total intracranial volume (TIV) and age at scan (Supplementary figure 1B).

### Supplementary analysis

In addition to this, we conducted a latent analysis model where we treated inflammation as an unobserved (“latent”) factor that manifests through IL-6, CRP and GlycA. Each biomarker was modelled as the latent inflammation factor multiplied by a factor loading—quantifying how strongly it reflects inflammation—plus a residual term capturing marker-specific noise. By fitting this simple CFA/SEM model, we obtain a single, more reliable inflammation score for each individual, which we then use in downstream analyses instead of juggling three correlated biomarkers. The results were not part of the main. The latent variable model was initially built as a minimally confounder-adjusted model but then constructed with full adjustments for confounders. For the model with a latent variable, a number of fit indices were then explored to test model adequacy, more specifically the Chi-square test, root mean square error of approximation (RMSEA) and the comparative fit index (CFI).

## Results

The sample consisted of 216 women with glycaemic measures at age 60-64 and neuroimaging data. A flowchart of the participants is presented in Figure 1 and sample characteristics are shown in Table 1.

**Figure 1:**
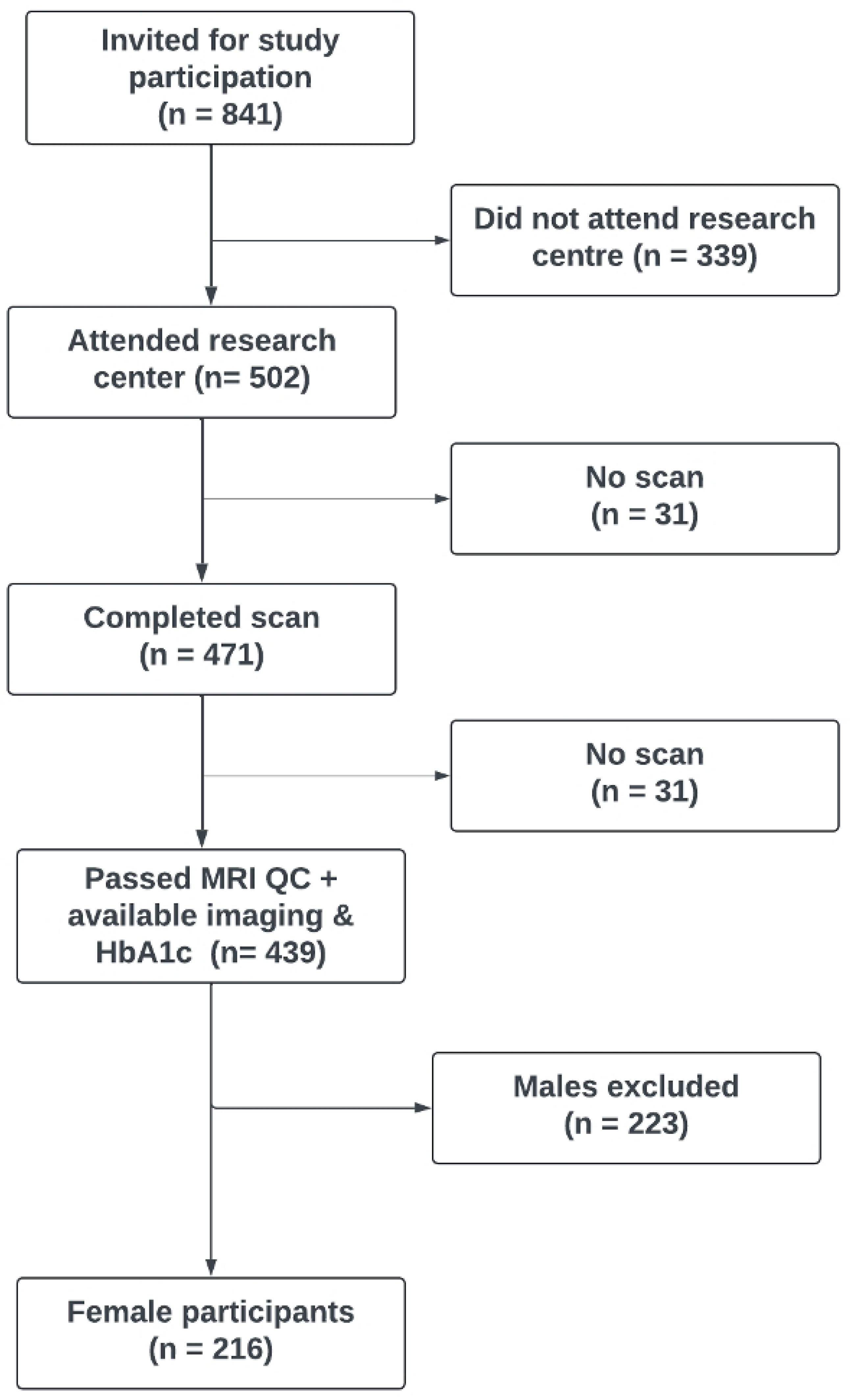
Flowchart providing an overview of Insight 46 recruitment of National Survey of Health and Development participants who undertook imaging and were part of my study. To be considered in this study, participants had to have available volumetric imaging data, HbA_1c_ data at age 60-64 and be a female. This amounted to 216 participants being included in the study.

**Table 1:**
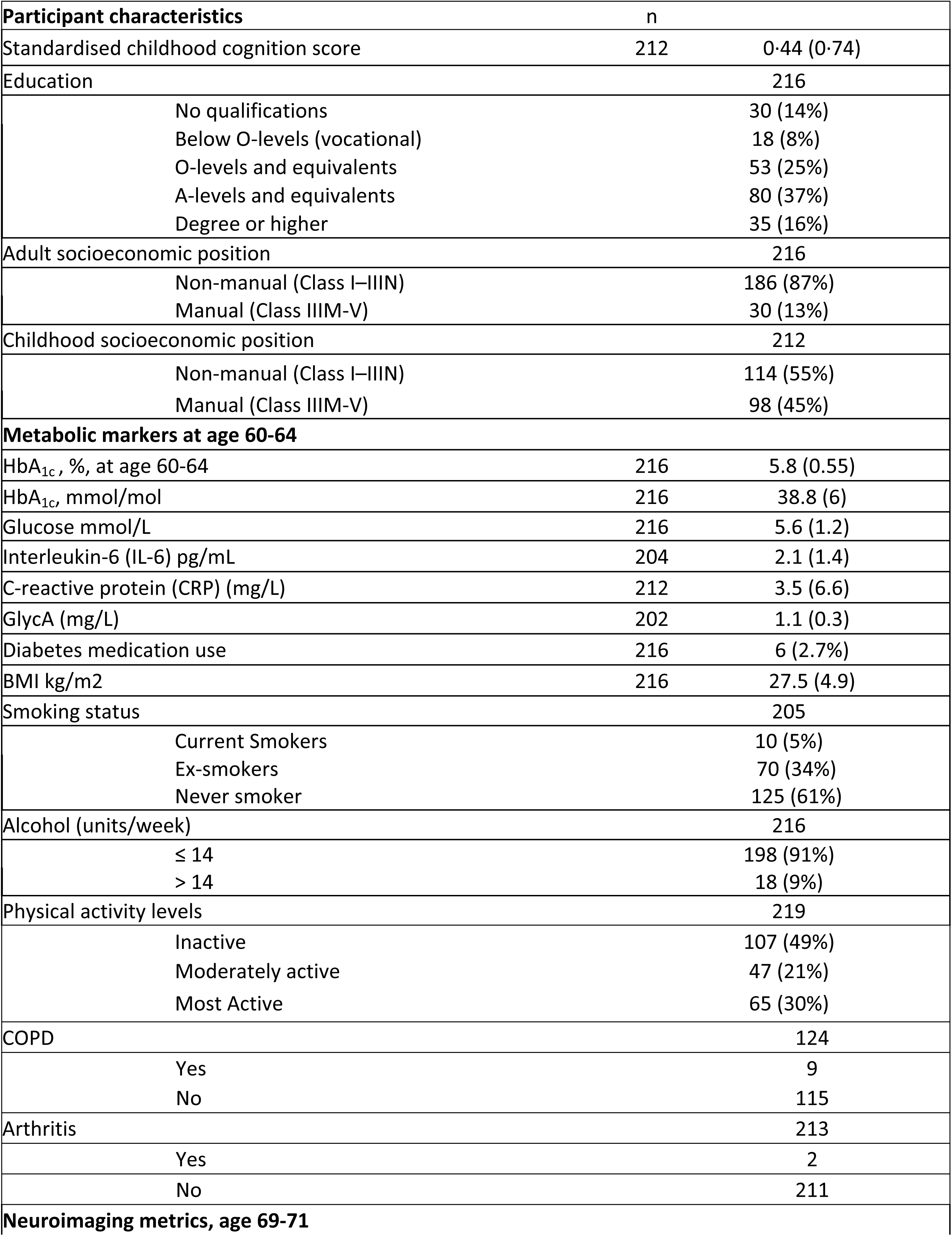
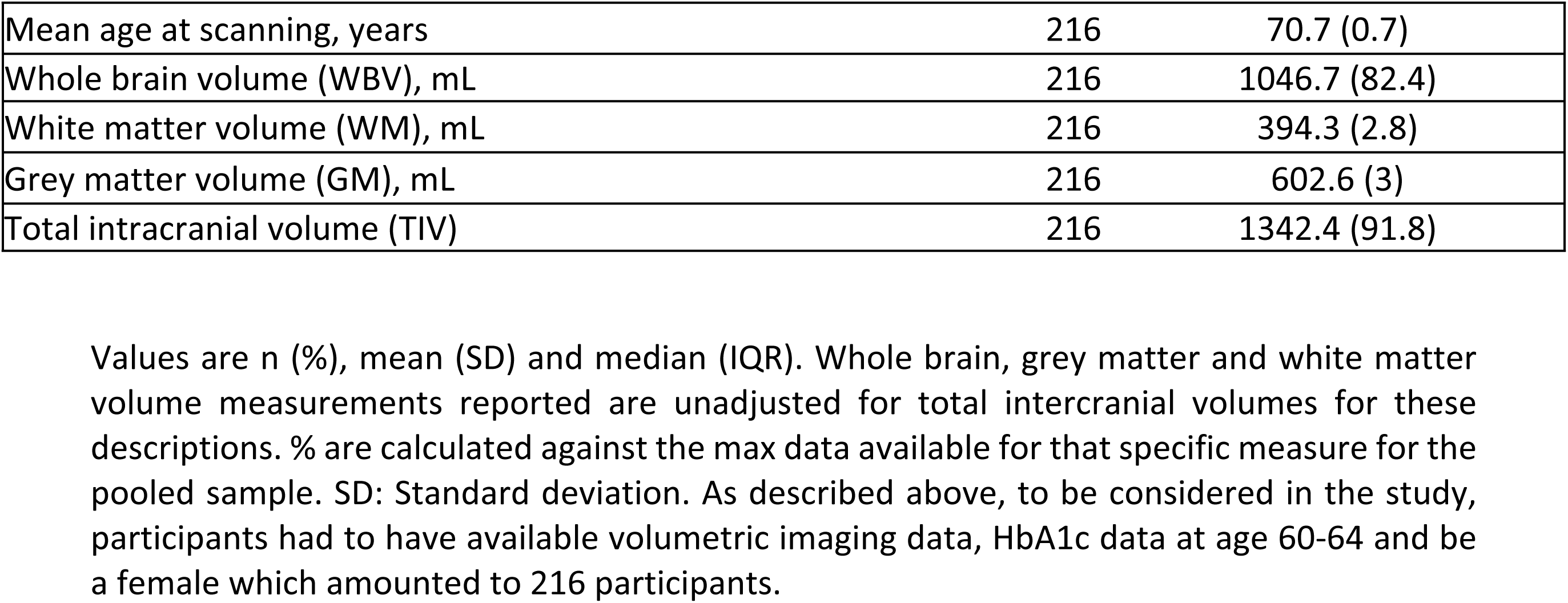
Sample characteristics for the participants considered in the analysis (max n = 216)

For IL-6 analyses, 10 participants with IL-6 levels above the maximum level reliably quantified by the current methods (10pg/mL) were excluded.

### Associations between inflammatory markers

The correlation matrix shows weak associations between the different inflammatory markers. The associations were strongest between CRP and IL-6 and least strong between GlycA and CRP. There were positive relationships between IL6, glycA and CRP. The relationship between IL6 and CRP was the strongest (r=0.4, p <0.001), followed by the association between glycA and CRP (r=0.2, p=0.004) and GlycA and IL6 (r=0.1, p=0.01).

#### Path analysis of HbA_1c_ and brain structure

As discussed in the methods, the results presented here are the path analysis for the fully confounder-adjusted models.

#### Whole brain volumes

There was a total effect of higher HbA1c at age 60-64 on lower WBV at age 69-71 for all the models (see Figure 2).

**Figure 2:**
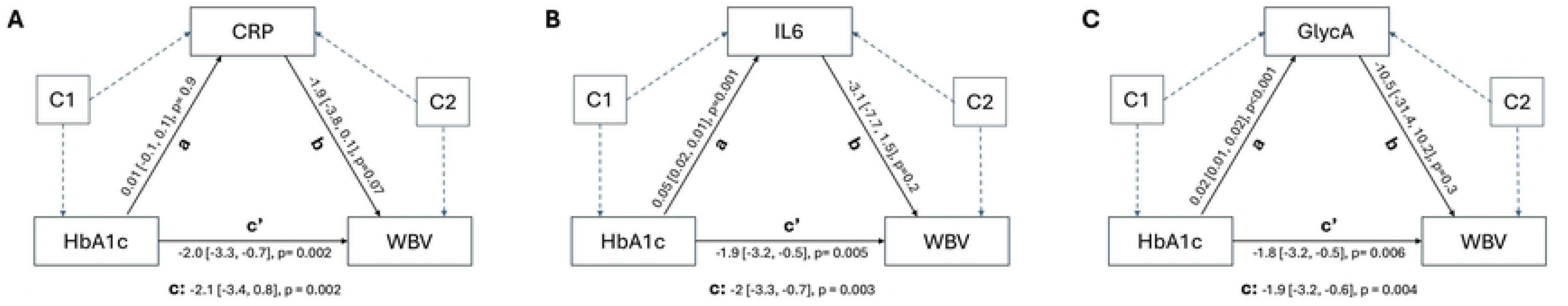
Mediation model path of HbA1c (exposure), inflammatory marker (mediator) and outcome (whole brain volumes). Model A considers CRP as the mediator. Model B considers IL-6 as the mediator. Model C considers GlycA as the mediator. As seen in each model: path **a** is the exposure → mediator link adjusted for exposure–mediator confounders path **b** is the mediator → outcome link adjusted for mediator-outcome confounders. path **c** is the total exposure → outcome effect before including the mediator. path **c′** is the direct exposure → outcome effect after including the mediator. Point estimates, 95 % confidence intervals, and *p*-values are shown for every path, with individual covariates collapsed into the two blocks **C1** (exposure-mediator confounders) and **C2** (mediator outcome associations) to aid visual clarity. Exposure-mediator confounders: Age, socioeconomic status, smoking, alcohol, body mass index and education Mediator-outcome confounders: Age, socioeconomic status, smoking, alcohol, body mass index and education chronic obstructive pulmonary dysfunction and arthritis.

In the mediation model, the link between HbA1c and whole-brain volume remained essentially a direct one, even after accounting for CRP, IL-6 and GlycA. There was no indication that any of those inflammatory markers carried part of the effect between HbA1c and brain volume. Separately, higher HbA1c was associated with elevations in IL-6 and GlycA, but it showed no meaningful relationship with CRP.

#### Grey matter volumes

Across all three inflammatory-marker models (CRP, IL-6, GlycA), higher HbA₁c was associated with reduced gray-matter volume, with total effect estimates of approximately (see figure 3). When CRP, IL-6 or GlycA were added to the model, the HbA₁c→GM relationship remained essentially unchanged, and none of the markers mediated this relationship (indirect effects very close to zero, all p>0.6) [see figure 3]. Finally, HbA₁c was itself positively related to IL-6 and GlycA (p≤0.005) but showed no meaningful link with CRP (p≈0.9).

**Figure 3:**
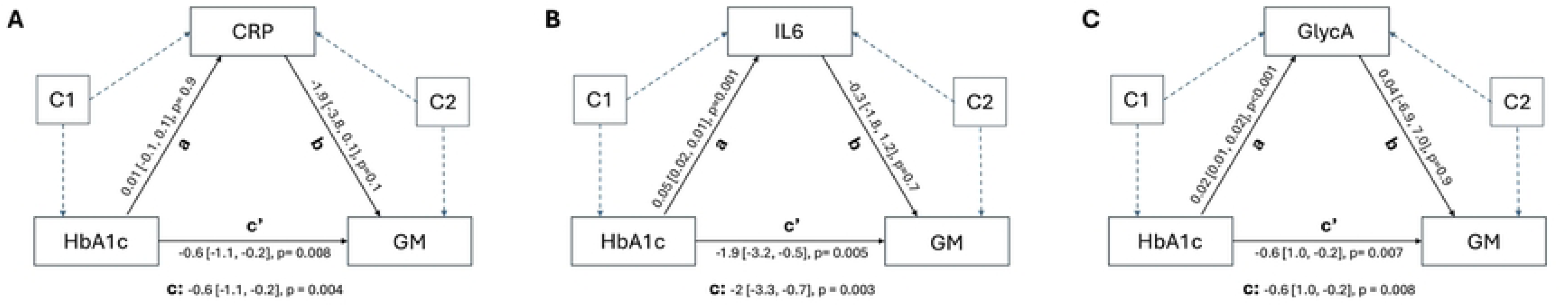
Mediation model path of HbA1c (exposure), inflammatory marker (mediator) and outcome (gray matter volumes). Model A considers CRP as the mediator. Model B considers IL-6 as the mediator. Model C considers GlycA as the mediator. As seen in each model: path **a** is the exposure → mediator link adjusted for exposure–mediator confounders path **b** is the mediator → outcome link adjusted for mediator-outcome confounders. path **c** is the total exposure → outcome effect before including the mediator. path **c′** is the direct exposure → outcome effect after including the mediator. Point estimates, 95 % confidence intervals, and *p*-values are shown for every path, with individual covariates collapsed into the two blocks **C1** (exposure-mediator confounders) and **C2** (mediator outcome associations) to aid visual clarity. Exposure-mediator confounders: Age, socioeconomic status, smoking, alcohol, body mass index and education Mediator-outcome confounders: Age, socioeconomic status, smoking, alcohol, body mass index and education chronic obstructive pulmonary dysfunction and arthritis.

#### White matter volumes

Since there was no total effect of HbA_1c_ on WM volumes, the results for these analyses were not decomposed into direct and indirect effects. For completeness, the tables are shown in [Figure 4]

**Figure 4:**
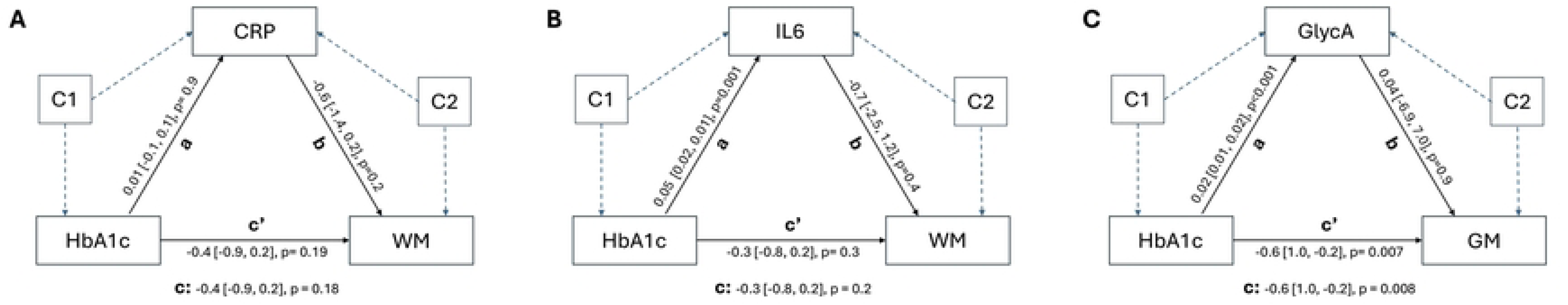
Mediation model path of HbA1c (exposure), inflammatory marker (mediator) and outcome (white matter volumes). Model A considers CRP as the mediator. Model B considers IL-6 as the mediator. Model C considers GlycA as the mediator. As seen in each model: path **a** is the exposure → mediator link adjusted for exposure–mediator confounders path **b** is the mediator → outcome link adjusted for mediator-outcome confounders. path **c** is the total exposure → outcome effect before including the mediator. path **c′** is the direct exposure → outcome effect after including the mediator. Point estimates, 95 % confidence intervals, and *p*-values are shown for every path, with individual covariates collapsed into the two blocks **C1** (exposure-mediator confounders) and **C2** (mediator outcome associations) to aid visual clarity. Exposure-mediator confounders: Age, socioeconomic status, smoking, alcohol, body mass index and education Mediator-outcome confounders: Age, socioeconomic status, smoking, alcohol, body mass index and education chronic obstructive pulmonary dysfunction and arthritis.

#### Glucose and brain structure

The findings for glucose on WBV, GM and WM volumes were similar to those for HbA_1c_ (Table 2). For WBV, GM and WM volumes, there was a total effect of glucose on WBV for CRP, IL-6, and GlycA models, with direct effects observed across these markers. However, there was no evidence of indirect effects through the inflammatory pathways. The exposure-mediator pathway was not statistically significant for CRP but was for IL-6 and GlycA. There was no evidence for the presence of a mediator-outcome pathway for any of the inflammatory markers. Overall, the trends observed for glucose were consistent with those seen for HbA_1c_, suggesting the role of both glycaemic measures do not involve indirect inflammatory pathways (Table 2).

**Table 2:** Path analysis of the fully confounder-adjusted models of the fasting glucose-brain associations (whole brain, grey matter and white matter volumes) via inflammation (c-reactive protein, interleukin-6 and glycoprotein-A). The table presents the β coefficients, confidence intervals and p values. The models were adjusted for TIV, age at scan, socioeconomic position, education, alcohol, smoking, physical activity, BMI, arthritis and COPD.

| Whole brain volumes (WBV) |  |  |  |  |  |  |  |  |  |  |  |  |  |
| --- | --- | --- | --- | --- | --- | --- | --- | --- | --- | --- | --- | --- | --- |
|  |  | c-reactive protein (CRP) |  |  |  | interleukin-6 (IL-6) |  |  |  | glycoprotein-A (GlycA) |  |  |  |
| | Path | $\beta$ | 95% CI | | p | $\beta$ | 95% CI | | p | $\beta$ | 95% CI | | p |
| Total effect | c | -6.5 | -11.4 | -1.6 | 0.009 | -6.5 | -11.4 | -1.6 | 0.01 | -6.4 | -11.3 | -1.5 | 0.01 |
| Direct effect | c' | -6.4 | -11.3 | -1.5 | 0.01 | -6.1 | -11.0 | -1.2 | 0.01 | -6.0 | -11.0 | -1.0 | 0.02 |
| Indirect effect |  | -0.1 | -0.8 | 0.5 | 0.7 | -0.4 | -1.1 | 0.3 | 0.3 | -0.4 | -1.5 | 0.6 | 0.4 |
| Exposure-mediator | a | 0.1 | -0.3 | 0.5 | 0.7 | 0.1 | -0.04 | 0.3 | 0.1 | 0.05 | 0.01 | 0.1 | 0.005 |
| Mediator-outcome | b | -2.0 | -3.5 | 0.4 | 0.1 | -3.4 | -8.1 | 1.3 | 0.1 | -9.4 | -30.6 | 11.6 | 0.38 |
| Grey matter volumes (GM) |  |  |  |  |  |  |  |  |  |  |  |  |  |
|  |  | c-reactive protein (CRP) |  |  |  | interleukin-6 (IL-6) |  |  |  | glycoprotein-A (GlycA) |  |  |  |
| | Path | $\beta$ | 95% CI | | p | $\beta$ | 95% CI | | p | $\beta$ | 95% CI | | p |
| Total effect | c | -1.7 | -3.4 | -0.09 | 0.04 | -1.7 | -3.4 | -0.05 | 0.04 | -1.7 | -3.4 | -0.04 | 0.04 |
| Direct effect | c' | -1.7 | -3.4 | -0.06 | 0.04 | -1.6 | -3.3 | -0.04 | 0.05 | -1.7 | -3.4 | -0.02 | 0.04 |
| Indirect effect |  | -0.03 | -0.2 | 0.1 | 0.7 | -0.05 | -0.2 | 0.1 | 0.6 | -0.001 | -0.3 | 0.3 | 0.9 |
| Exposure-mediator | a | 0.1 | -0.3 | 0.5 | 0.7 | 0.1 | -0.04 | 0.3 | 0.1 | 0.05 | 0.01 | 0.08 | 0.005 |
| Mediator-outcome | b | -0.4 | -1.0 | 0.2 | 0.2 | -0.4 | -1.9 | 1.0 | 0.5 | -0.03 | -7.1 | 7.0 | 0.9 |

| White matter volumes (WM) |  |  |  |  |  |  |  |  |  |  |  |  |  |
| --- | --- | --- | --- | --- | --- | --- | --- | --- | --- | --- | --- | --- | --- |
|  |  | c-reactive protein (CRP) |  |  |  | interleukin-6 (IL-6) |  |  |  | glycoprotein-A (GlycA) |  |  |  |
|  | Path | β | 95% CI |  | p | β | 95% CI |  | p | β | 95% CI |  | p |
| Total effect | c | -2.3 | -4.3 | -0.3 | 0.02 | -2.2 | -4.2 | -0.2 | 0.03 | -2.2 | -4.2 | -0.2 | 0.03 |
| Direct effect | c' | -2.2 | -4.2 | -0.3 | 0.03 | -2.2 | -4.2 | -0.1 | 0.04 | -2.0 | -4.0 | -0.3 | 0.05 |
| Indirect effect |  | -0.05 | -0.03 | 0.2 | 0.7 | -0.01 | -0.3 | 0.2 | 0.5 | -0.2 | -0.7 | 0.2 | 0.3 |
| Exposure-mediator | a | 0.1 | -0.3 | 0.5 | 0.7 | 0.1 | -0.04 | 0.3 | 0.1 | 0.05 | 0.01 | 0.08 | 0.006 |
| Mediator-outcome | b | -0.6 | -1.4 | 0.2 | 0.1 | -0.7 | -2.5 | 1.1 | 0.4 | -5.3 | -13.7 | 3.1 | 0.2 |

#### Latent variable analysis

For the latent model of inflammatory markers, there was evidence of a total and direct effect of with HbA_1c_ and glucose on WBV, but no total or direct effect on GM or WM volumes (Supplementary Table 1). There was no evidence of indirect effects observed between HbA_1c_ or glucose on WBV, GM, or WM volumes through the inflammatory pathway.

## Discussion

### Summary of findings

The aim of this mediation analysis was to gain better insight into an important potential mechanism linking hyperglycaemia with smaller brain volumes (Fatih et al, 2022). The path analyses demonstrated that there was: 1) a total effect of glycaemic markers on smaller brain structure, 2) hyperglycaemia was mostly associated with higher systemic inflammation (for IL-6 and GlycA) but 3) there we found no association between the systemic inflammatory markers and volumetric brain measures. Hence overall there was no indirect effect of glycaemia on these brain outcomes via a systemic inflammation pathway.

Considering the inflammatory markers as a latent variable for inflammation had little influence on the results nor did considering fasting glucose measure as the exposure of any analyses. Overall, the findings suggested that while both high glucose and increased inflammation are related, systemic inflammation, as measured by serum biomarkers, does not mediate the relationship between glucose levels and smaller brain volume in this sample of women.

### Specific findings and concordance with the literature

The study was motivated by previous research suggesting that individuals with T2D show elevated levels of inflammatory markers compared to healthy controls (regardless of disease duration) (Okdahl et al., 2022). Similarly, women with T2D showed higher inflammation TNF-α, IL-6 and CRP compared to women without the condition (Hu et al., 2004; Liu, 2007). There is also evidence that inflammatory markers are associated with poorer brain health and that cytokines accumulate at different rates in AD patients compared with healthy control subjects (Lue, 2001; Marsland et al., 2008; Walker et al., 2017).

Our observation that poorer glycaemia was associated with higher inflammation, is in line with previous evidence in a mixed sample of individuals with T2D and healthy controls.(Gautam et al., 2023) This is consistent with the known effect of hyperglycaemia on NF-κB–dependent inflammatory cytokine production and other mechanisms linked to the activation of inflammatory pathways(Gupta et al., 2023; Shukla et al., 2017).

The evidence linking inflammation to brain health is less consistent. Previous analyses from population-based studies have revealed mixed findings in relation to the association between markers of systemic inflammation and brain health outcomes. For example, recent findings from the Atherosclerosis Risk in Communities (ARIC) study failed to find an association between inflammatory markers and WBV (Walker et al., 2017) consistent with our findings. In contrast, the Framingham study found that most, but not all, of the inflammatory markers they considered were associated with lower total brain volume (Jefferson et al., 2007).

Discrepancy in the results to the Framingham study may be explained by the different methodological approaches taken. Firstly, Framingham considered a mixed-sex sample and, notably, reported stronger inverse inflammation-brain association in men, with the corresponding slopes in women were smaller and often non-significant. For example, the association between IL-6 and their brain outcome ranged in the null in women (akin to ours) but was robust in men.

In our analysis, we only considered women and had a narrower biomarker scope (only available data for CRP, IL-6 and GlycA) comparatively to Framingham who assayed ten cytokines, allowing robust signals from markers such as OPG and TNF-α to emerge. Despite this, our findings for IL-6 were largely consistent with those of their study. Framingham also considered a head-size-normalised total cerebral brain volume (TCBV) measured in very close temporal proximity to their biomarker (biomarker and scan on the same visit). We linked our inflammatory biomarkers measured at ages 60–64 to raw whole-, grey- and white-matter volumes ten years later, then fitted mediation model. Taken together—our women-only focus, the narrow biomarker panel, the wider (ten-year) interval between biomarker assessment and MRI, the use of absolute brain-volume measures, and different confounder adjustment may explain differences in the findings. Since their findings were less notable in their women cohort (and at times consistent with ours e.g. IL-6), it may be argued that the absence of a significant effect in NSHD is compatible with, rather than contradictory to, the modest negative trend reported for women in the Framingham cohort.

There remains an important question, that is whether markers of systemic inflammation can give us a robust insight into brain inflammation. Some previous studies have found that systemic inflammation, particularly through infections, can influence neuroinflammatory processes, but the interplay is complex (Lassarén et al., 2021). Systemic IL-6 and other cytokines contribute to immune changes in the brain, yet these effects are modulated by local brain mechanisms and do not necessarily follow a linear systemic-to-brain pathway.

For instance, systemic infections can elevate arterial cytokine levels (e.g., IL-6, IL-8, IL-10) while decreasing certain cytokines in the brain’s extracellular fluid (Lassarén et al., 2021). This intricate relationship highlights the challenges of relying solely on systemic inflammatory markers to understand what may be happening in the brain and underscores the need for more precise biomarkers to elucidate the impact of neuroinflammation on brain health and neurological disorders.

Interestingly, we observed an association between glycaemia and GlycA and IL-6 not CRP. Mechanistically, studies have shown that IL-6 and CRP are closely related, with IL-6 being the major factor that triggers the hepatic synthesis of CRP (Boras et al., 2014; Sproston & Ashworth, 2018). However, although inflammatory markers are usually linked, recent studies have found them to diverge in certain contexts. For example, differing concentrations of IL-6 and CRP have been found in relation to HRT use (Bermudez et al., 2002). Similarly, divergent levels of these inflammatory markers have been found in relation to other clinical factors such as alcohol use and exercise (Bermudez et al., 2002). Thus, although CRP and IL-6 are biologically linked, their levels can diverge under certain conditions which may account for the weak correlation observed in this analysis.

Considering inflammation as a latent variable offered no additional insight into these associations (see supplementary table 1). When designing this study, we made the decision to construct a latent variable for inflammation using IL-6, CRP and GlycA. The rationale was that combining multiple measures may give a more comprehensive insight into systemic inflammation by capturing the common variance amongst them and reducing measurement error. However, the general lack of correlation between the inflammatory variables may have limited the potential utility in line with the fit indices suggesting some inconsistency in terms of the model’s adequacy.

It is possible that the relationship between hyperglycaemia and brain health is mediated by factors other than inflammation. While we found no evidence that systemic inflammation directly mediates the relationship between HbA_1c_ and WBV, other possible mediators to consider may be oxidative stress, small vessel disease and CVD (Gupta et al., 2023). Other factors that may be particularly important around late midlife include hormonal health such as oestrogen and its neuroprotective effect reducing during the perimenopause and the post menopause. This change in hormonal health can influence insulin sensitivity and glucose metabolism (Kautzky-Willer et al., 2016) as well as brain structure and function in women (Mosconi et al., 2021) Additionally, non-biological factors in social roles and responsibilities, including caregiving duties and family responsibilities, may mediate these sex-specific associations found in women by imposing chronic stress and time constraints, which might affect brain health. Future research should consider important biological and social factors specific to women to achieve a comprehensive understanding of the underlying mechanisms of the glycaemia-brain health pathways.

In this study, we considered that hyperglycaemia precedes inflammation. This is based on previous studies suggesting that hyperglycaemia and abnormal glucose metabolism can result in the formation of advanced glycation end products (AGES) with reactive oxygen species (ROS)triggers pathways that regulate the inflammatory response resulting in the increase of pro-inflammatory cytokines such as IL-6 (Gupta et al., 2023). However, these cytokines produced by adipose tissue and macrophages may also result in a state of IR thus contributing to the pathophysiology of T2D (Pradhan, 2001) This was found even when adjustments were made for inflammatory-related confounders (e.g., smoking, exercise, and BMI). In an attempt to reduce the likelihood of infection-driven hyperglycaemia, participants with an IL-6 value of > 10 pg/mL were excluded prior to my analyses.

### Strengths and weaknesses

The study has multiple strengths. First, it considered multiple markers of inflammation. Few population-based studies have had the availability of multiple inflammatory markers. The confounder data includes those for the exposure-mediator and the mediator-outcome paths, which are known problems with causal mediation studies (Cole & Hernán, 2002).

Furthermore, an important strength of this study is that it considered samples of women of the same age. The homogeneity of age ensured age-specific brain changes did not confound the results, providing a clearer picture of the relationship between HbA_1c_ and brain health.

Since inflammatory markers were not associated with the brain imaging measures considered, further studies should consider their associations to more subtle markers of brain pathology such as those of microstructural diffusion metrics and enlarged periventricular spaces. This will provide an insight into the impact of diabetes-related inflammation on early-stage small vessel disease.

It is worth acknowledging the potential for reverse causation, where changes in brain volume could influence metabolic parameters, rather than the reverse. This highlights the need for caution in interpreting the directionality of the observed relationships.

Additionally, other unmeasured variables not considered in this analysis, such as genetic factors or specific dietary components, might influence the relationships explored.

## Conclusions

These findings reveal that for women of this birth cohort, the relationship between glycaemia in midlife and later life smaller brains was not mediated by systemic inflammation as measured by selected blood markers. As per the findings we reported in our previous paper, poorer glycaemia is directly associated with smaller brains but there was no indirect path of this relationship through inflammation. Future studies could investigate these associations in a sample such as UK Biobank to examine the causal the nature of these relationships through techniques such as Mendellian Randomisation.

## Data availability

Anonymized data will be shared by request from qualified investigators (skylark.ucl.ac.uk/NSHD/doku.php).

## Data Availability

Data from the MRC National Survey of Health and Development (NSHD), including the Insight 46 sub-study, are available to bona fide researchers on request from the Unit for Lifelong Health and Ageing at UCL application details are provided on the NSHD data-sharing webpage.

https://skylark.ucl.ac.uk/NSHD/forms/data-access-request-form/

## Acknowledgements

We are very grateful to those study members who helped in the design of the study through focus groups, and to the participants both for their contributions to Insight 46 and for their commitments to research over the last seven decades.

## Author contribution

All authors made substantial contributions to this manuscript. NF conceptualised and formally analysed the data under the supervision of NC, AH and SNJ and with statistical advice from RS. MR and JMS acquired funding. SNJ, JB, CS, MR, DC, IM, JMS were responsible for acquisition of the data. NC, VG, AH, SJ were crucial for their insight to help supervise the project. NF prepared the original. All authors reviewed, left commentary review then revised and approved the manuscript.

## Supporting Information Captions

**Supplementary Figure 1:**
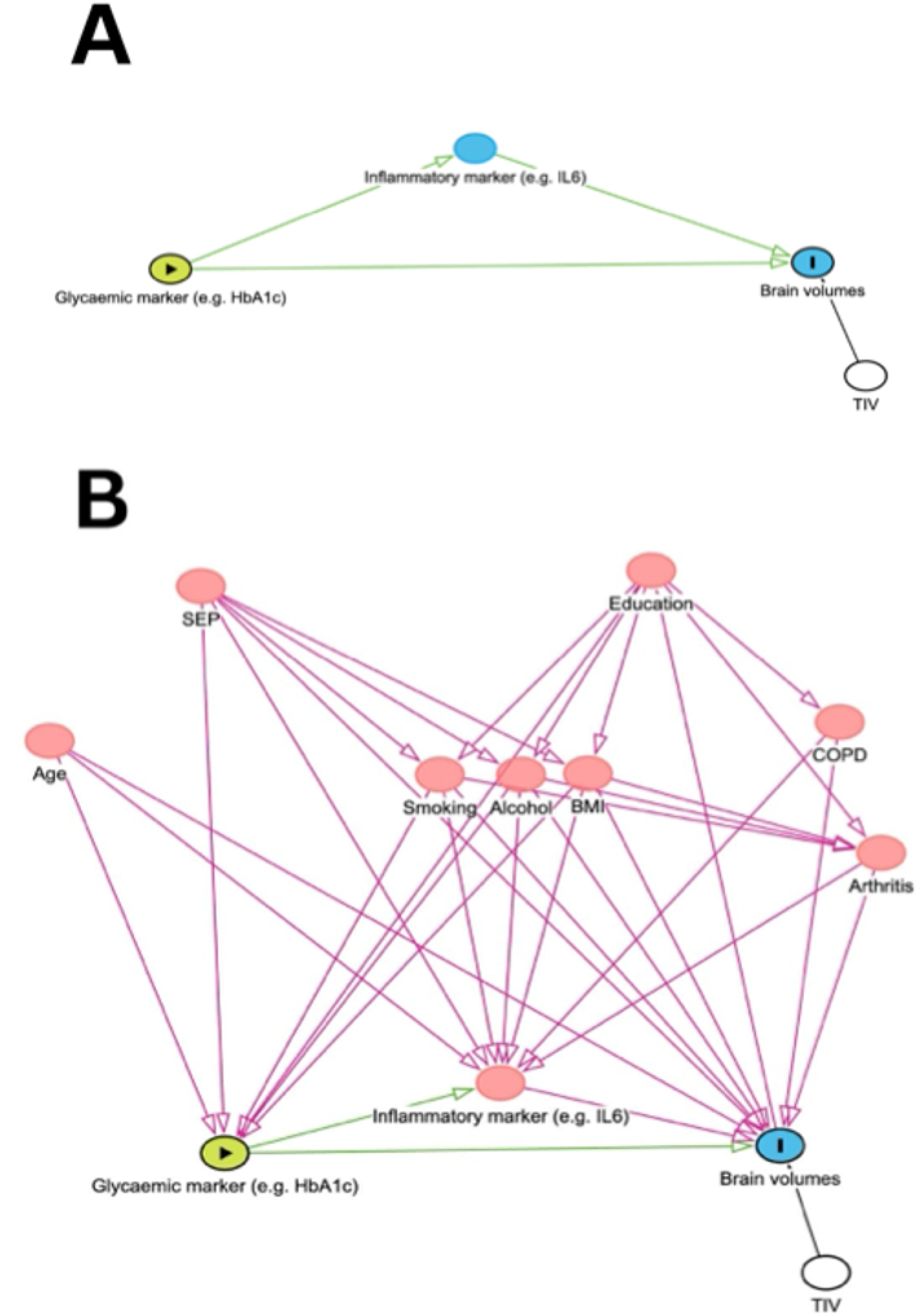
Path analysis models conducted in the analyses A) Simple mediation model as a first step of model building. B) Fully confounder-adjusted model. Adjustments were made for both exposure-mediator and mediator-outcome relationships.

**Supplementary Figure 2:**
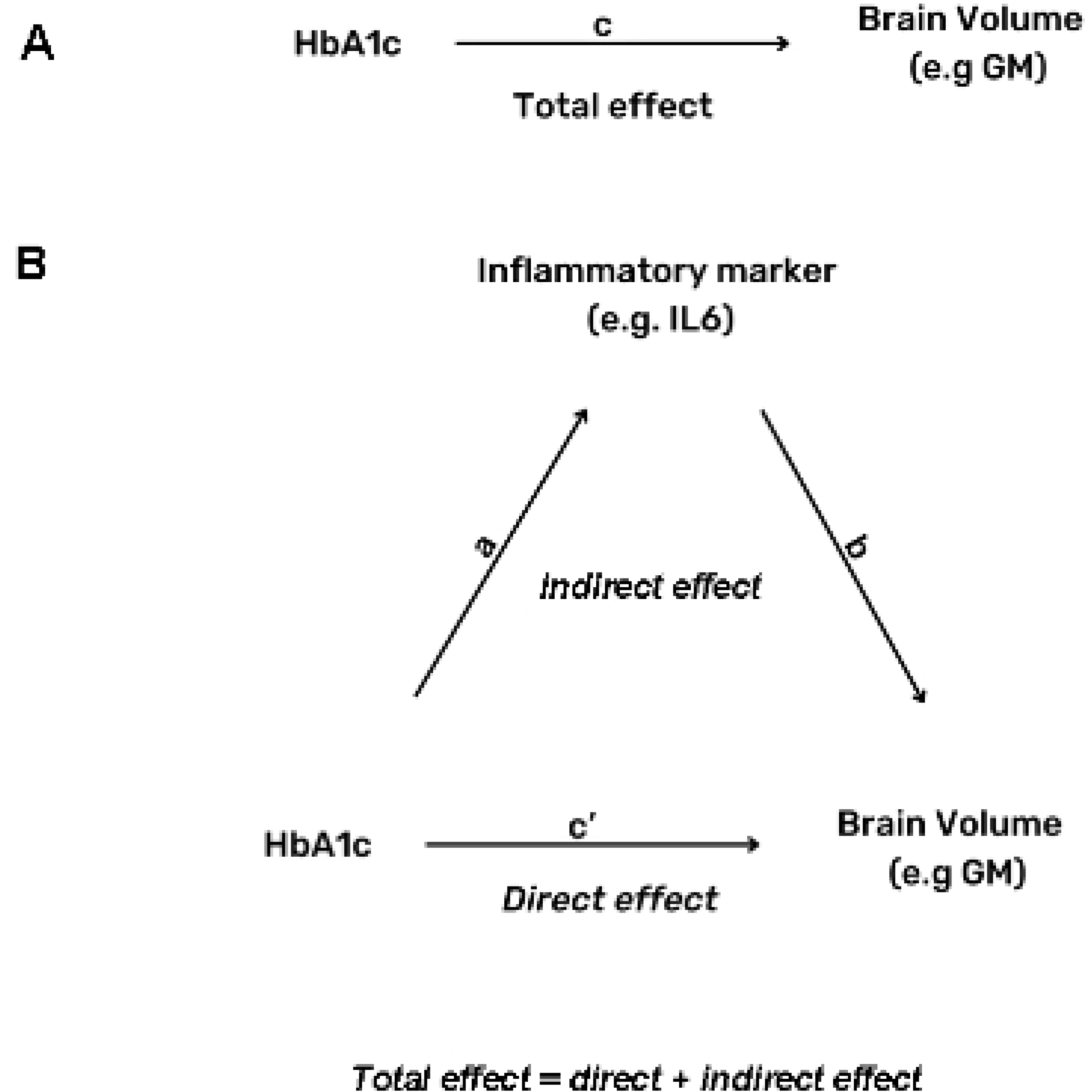
A theoretical analysis of the mediation models considered. A) shows the assumed (total) effect, c. B) the total effect decomposed into a direct, c’ and indirect (mediated via a and b) effect.

Supplementary table 1: Path analysis output of the latent model for the HbA_1c_ and glucose-brain associations (whole brain, grey matter and white matter volumes) via the latent variable of inflammation (constructed using c-reactive protein, interleukin-6 and glycoprotein-a). The total effects, direct, indirect effects, exposure-mediator are presented. β, Cl and p-values are presented.

